# Waning of PCV13 vaccine-induced antibody levels within the first year of life, using a 3+0 schedule: an observational population-level serosurveillance study among children under 5 years old in Blantyre, Malawi

**DOI:** 10.1101/2022.04.21.22272868

**Authors:** Todd D Swarthout, Marc Y R Henrion, Deus Thindwa, James E Meiring, Maurice Mbewe, Comfort Brown, Jacquline Msefula, Brewster Moyo, Andrew A Mataya, Susanne Barnaba, Emma Pearce, Melita Gordon, David Goldblatt, Neil French, Robert S. Heyderman

## Abstract

**Background:** Pneumococcal conjugate vaccines (PCVs) induce serotype-specific IgG antibody, effectively reducing vaccine-serotype (VT) carriage and invasive pneumococcal disease (IPD). IgG production wanes approximately 1 month after vaccination in absence of serotype-specific exposure. With uncertainty around correlate of protection (CoP) estimates and with persistent VT carriage and VT-IPD following PCV13 introduction, we undertook population-level immunogenicity profiling among children <5 years in Blantyre, Malawi.

**Methods:** For 638 children, capsule-specific IgG to PCV13 VTs, two non-VTs, and IgG to three pneumococcal proteins were measured using an enzyme-linked immunosorbent assay and a direct-binding electrochemiluminescence-based multiplex assay. A linear spline regression model estimated population-level, serotype-specific immunogenicity profiles. A linear regression model was used to validate putative CoPs.

**Findings:** Immunogenicity profiles revealed a consistent pattern among VTs except serotype 3: a vaccine-induced IgG peak followed by waning to a nadir and subsequent increase in titre. For serotype 3 there was no apparent vaccine-induced increase. Heterogeneity in parameters included age range at post-vaccination-nadir (11·2 [19F, 23F] to 27·3 [7F] months). Titres dropped below IPD CoPs among 9 VTs and below carriage CoPs for 10 VTs. Study data estimated a range of carriage CoPs (0·50μg/mL to 2·5μg/mL). Increasing antibody among older children and seroincident events were consistent with ongoing VT exposure.

**Interpretation:** A 3+0 PCV13 schedule with high uptake has not led to sustained population-level antibody immunity beyond the first year of life. Indeed, post-vaccine antibody concentrations dropped below putative CoPs for several VTs, potentially contributing to persistent VT carriage and residual VT-IPD in Malawi and other similar settings.

**Funding:** Bill & Melinda Gates Foundation, Wellcome UK, and National Institute for Health & Care Research.

## INTRODUCTION

The encapsulated bacteria *Streptococcus pneumoniae* (the pneumococcus) is responsible for approximately 300 000 deaths worldwide, with one-third of these occurring among children under 5 years of age and with the greatest burden in low- and middle-income countries (LMICs).^1^ *S. pneumoniae* has almost 100 capsule serotypes, with 20–30 accounting for most invasive pneumococcal disease (IPD). Nasopharyngeal carriage of the pneumococcus in the upper airway is a prerequisite for disease but is also a key process in the development of natural immunity.^2^ Immunocompromised individuals, infants and older adults are at greater risk of IPD, with LMICs bearing the greatest burden.^3,4^

Pneumococcal conjugate vaccines (PCVs), currently including 10 (PCV10) or 13 (PCV13) capsular polysaccharides, have been introduced into routine infant immunisation programmes across the world. These have been effective in reducing nasopharyngeal carriage prevalence and IPD incidence among both vaccinated (direct protection) and unvaccinated (indirect protection) children and adults. PCVs elicit protection against disease in infants through induction of serotype-specific opsonophagocytic antibodies.^5^ Immunoglobulin G (IgG) antibody production induced by PCV peaks approximately 1 month after vaccination and wanes thereafter in the absence of serotype-specific natural exposure.^6^ A pooled serum IgG correlate of protection (CoP) against IPD of 0·35μg/mL was derived from a meta-analysis of three efficacy trials.^7^ Although use of 0·35μg/mL as an aggregate CoP has enabled the licensing of new PCVs, Andrews and colleagues have subsequently reported serotype-specific CoPs that vary widely, with some higher (1, 3, 7F, 19A, 19F) and others lower (6A, 6B, 18C, and 23F) than 0·35μg/mL (supplementary table 1).^8^

A CoP against pneumococcal carriage has been more problematic to derive, in part because of the complexity of interactions between the pneumococcus and the human mucosal immune system.^9^ Here, serotype-specific effector mechanisms likely include both circulating serum IgG and locally produced IgA and IgG in the context of T-cell immunity to sub-capsular protein antigens, cellular and soluble innate immunity.^10,11^ To overcome considerable heterogeneity, Voysey and colleagues used data from multiple vaccine immunogenicity trials to calculate serotype-specific carriage CoPs, reporting estimates that are i) higher for carriage than for IPD and ii) serotype-specific, with 6B among the lowest (0·50μg/mL) and 19F among the highest (2·54μg/mL; supplementary table 1).^12,13^ Importantly, they also found that protective correlates were, on average, two times higher in LMICs than in high income countries.

Malawi introduced PCV13 into the national Expanded Programme of Immunisations (EPI) in 2011, using a 3+0 schedule (one dose at each 6, 10 and 14 weeks of age) with a catch-up campaign limited to children aged under-1 year. Field studies report high vaccine coverage, exceeding 90%.^14^ Though we have shown reduced VT carriage in Malawi, we saw persistent residual carriage of all 13 VTs among PCV13-vaccinated children up to 7 years after PCV13 introduction.^15^ Similar patterns have been seen in Kenya^16^ and The Gambia.^17^ We also showed that, despite a vaccine-attributable reduction in VT-IPD among vaccine-age-eligible children, there is persistent residual VT-IPD among both PCV-vaccinated and PCV-unvaccinated individuals.^18^ This suggests sub-optimal control of colonisation and onward transmission, as well as persistent vulnerability to VT-IPD, even among PCV13-vaccinated children.

We have suggested that a high force of infection in settings such as Malawi contributes to a short duration of PCV13-induced VT carriage control,^19^ hypothesising that rapid waning of vaccine-induced antibody resulting from the 3+0 schedule enables persistent VT carriage. To further evaluate the complex interaction between immunity (vaccine-induced and through natural exposure) and colonisation, we have undertaken population-based pneumococcal serology profiling and carriage surveys among children aged under-5-years in Blantyre, Malawi. We describe the serotype-specific IgG profiles among children aged under-5-years and have evaluated their likely impact on carriage acquisition and IPD among key PCV13 serotypes using candidate serological CoPs.

## METHODS

### Study setting and participants

Malawi is a low-income country in southern Africa. Blantyre (population 1·3 million) is the geographical centre of the country’s Southern Region. In 2011, Malawi adopted the WHO-recommended Option B+, whereby all HIV-infected pregnant or breastfeeding women commence lifelong antiretroviral therapy regardless of clinical or immunological stage, dramatically reducing mother-to-child transmission of HIV.

### Pneumococcal carriage

A series of rolling pneumococcal carriage surveys (the PCVPA Study) were implemented between 2015 and 2019 in Blantyre, randomly sampling healthy participants, including children 18 weeks to 7 years old (PCV13 vaccinated) and children 3–10 years old (age-ineligible to receive PCV13).^15^ Despite evidence of reduced VT carriage, we identified persistent residual carriage of all 13 VTs up to 7 years after PCV13 introduction.^15^ Supplementary Table 2 shows the VT carriage prevalence among children from the PCVPA Study.

### Serology

A random subset of samples was selected from a prospective population-based serosurvey undertaken in 2016–2018.^20,21^ The serosurvey was implemented in Ndirande, the same urban township in Blantyre where the PCVPA Study was implemented, though recruitment did not necessarily include the same children. Individuals were randomly selected from a population demographic census and invited to participate. Children’s vaccination status was obtained from patient-retained health books (known as health passports) or, if documentation was unavailable, from parents or guardians. All participants were followed up to provide a second linked sample approximately 3 months after the first sample. For the purposes of this evaluation, we randomly selected 556 primary samples, of which 82 had linked secondary samples. This sample size allowed for predicting the time, after vaccination, when 50% of the population would fall below an anti-capsular antibody level (0·35 & 1·0μg/ml) with a 95% precision of 8 weeks (ie, ±4 weeks around the point estimate). The dynamics of antibody decay was based on work by Ota and colleagues.^22^

### Plasma preparation

Venous EDTA samples were stored in cool boxes upon collection and transported to the study laboratory. Plasma was separated by centrifugation and stored at −80°C within 8 hours of collection.

### Antibody Measurement

Serological analysis was performed at the World Health Organisation (WHO) reference laboratory for pneumococcal serology, Great Ormond Street Institute of Child Health, University College London, UK. Plasma was stored at −70°C prior to assay for serotype-specific IgG to the 13 VT capsular polysaccharides (1, 3, 4, 5, 6A, 6B, 7F, 9V, 14, 18C, 19A, 19F and 23F). Plasma were assayed using the WHO reference enzyme-linked immunosorbent assay (ELISA) following adsorption with cell wall polysaccharide and 22F polysaccharide at a concentration of 10μg/mL as previously described.^23^ This assay is based on the original Wyeth assay used to generate the aggregate CoP of 0·35 µg/mL. The lower limit of assay quantification was 0·15µg/mL. Serum IgG to pneumococcal protein antigens were measured using a direct binding electrochemiluminescence-based multiplex assay (MesoScale Discovery (MSD; Maryland, USA) on customised antigen plates as previously described.^24^ Functional antibodies to 13 serotypes were measured in a multiplexed opsonophagocytic assay (OPA) as previously described.^25^ Values are expressed as an OPA titre, equivalent to the reciprocal of the serum dilution required to produce 50% killing of the relevant serotype.

### Statistical analysis

#### Linear spline regression models

The relationship between age and IgG is nonlinear and changes with age. To model the nonlinear relationship between age and IgG with generalised linear regression models and to perform statistical inference on the locations of the change points, we used linear splines. The regression models we developed are piecewise linear models, joined at specific values of the predictor variable. These locations, referred to as changepoints or knots, define the intervals over which the relationship between predictor and response is assumed to be linear. As a prerequisite, the fitted models needed to satisfy several requirements, including i) properly accounting for data values that are left censored (below the assay detection limit), ii) treating the knot locations as parameter values to be estimated during model fitting, iii) returning a fit for the geometric mean IgG concentration as a function of age, and iv) selecting the optimal number of changepoints. To achieve this, we fitted censored regression models with linear splines for the age predictor variable, treating knot locations as additional model parameters and manually optimised the resulting likelihood function. We used bootstrapping and the percentile method to derive confidence intervals for slope parameters and knot locations. Before model fitting, the IgG data were log-transformed so that the fitted arithmetic mean corresponded with the log of the geometric mean on the original data scale. For every serotype, we fitted models with k=0, 1, 2, 3 knots and then selected the optimal value for k using the Akaike Information Criterion. Implementation was done using R version 4·1·2 (R Foundation for Statistical Computing, Vienna, Austria). The modelling code is available on GitHub.^26^ Further details on the modelling methodology are provided in the Supplementary Material.

### Evaluating serological CoPs against carriage acquisition

Carriage CoPs are likely to vary by population, depending on several factors including the force of infection, crowding, nutritional status, and air pollution. Therefore, in addition to referencing previously reported CoPs for carriage, we used serotype-specific serological data from this study and previously reported carriage prevalence from the same population^15^ for further validation. Carriage prevalence was used as a proxy for transmission intensity. Carriage prevalence and median IgG titre were calculated for each age group: 0–2 months, 3–5 months, 6–11 months, and groupings covering every 12–month intervals until 5 years of age. Empirical CoPs were estimated using either of two possible approaches. The first approach involved aligning the carriage prevalence and serology data among recently vaccinated age groups before vaccine-induced IgG waned significantly (including at 3–6 months and 6–12 months of age); if there was no or low carriage prevalence in these age groups the reported serological titre was presumed to be higher than the CoP threshold. For the second approach, using the correlation between carriage prevalence and serology for all age groups under different transmission intensities, we used a linear regression between carriage prevalence and median serotype-specific IgG titre, defining the CoP as the predicted value from the fitted regression model at 0% prevalence if there was an association.

### Ethics approval and consent to participate

The pneumococcal carriage study protocol was approved by the Kamuzu University of Health Sciences (formerly College of Medicine, University of Malawi) Research and Ethics Committee and the Liverpool School of Tropical Medicine Research Ethics Committee. The serology component received ethics approval from the Oxford Tropical Research Ethics Committee, the Malawian National Health Sciences Research Committee, and the Kamuzu University of Health Sciences (formerly College of Medicine, University of Malawi) Research and Ethics Committee. Parents/guardians of child participants provided written informed consent, including consent for publication.

## RESULTS

### Recruitment

We evaluated 638 plasma samples for serotype-specific IgG titres among children aged under-5-years, including 556 primary samples and 82 unique secondary samples (each linked to one primary sample). Among participants, 52·0% were male, and the median age at primary sample collection was 1·9 years (range 42 days to 4·9 years; table 1). Secondary plasma samples were collected approximately 3 months (median 98 days, range 83–107) after the primary sample.

**Table 1.** Participant characteristics

| Age | n | Gender, male % | Household size, median (IQR) | Primary caregiver, secondary education completed, % | Primary caregiver, employed (yes), % |
| --- | --- | --- | --- | --- | --- |
| <3mo | 11 | 63.6 | 6 (4-9) | 50.0 | 100.0 |
| 3-5mo | 15 | 53.3 | 4 (4-5) | 40.0 | 100.0 |
| 6-11mo | 89 | 48.3 | 5 (4-6) | 43.2 | 96.6 |
| 1y | 188 | 51.1 | 5 (4-6) | 48.7 | 88.2 |
| 2y | 84 | 54.8 | 5 (4-6) | 40.3 | 91.7 |
| 3y | 78 | 42.3 | 5 (4-6) | 40.3 | 91.7 |
| 4y | 82 | 56.1 | 5 (4-7) | 40.4 | 88.7 |
| Total | 547<sup>1</sup> | 51.0 | 5 (4-6) | 43.7 | 91.2 |

<sup>1</sup>Analysis is based on available data from 547 of 556 primary samples. IQR=interquartile ratio, mo=months, y=years

The 556 primary samples were stratified to twenty 3-month age strata (eg, aged <3 months through to 57–59 months), with a median 21 samples (range 9 to 54) per stratum (supplementary table 4A and 4B). About two-thirds (67·2%) of the children had health passports (patient-retained health records). Based on either health passport or information from guardians, completion of 3-dose PCV13 vaccination was 98·1% with all children having received at least one dose. In comparing age strata, the distribution of participant characteristics was similar (table 1).

### Serological assay output

The number of serology results per serotype is shown in supplementary table 3. In brief, a range of 599 (serotype 3) to 637 (serotype 6A) samples provided serotype-specific IgG concentration data included in the analysis.

### Linear spline regression model

The population-level spline model output, based on 556 primary samples (figure 1 and table 2), shows a consistent pattern among all VTs, except serotype 3: There is a period among young infants with lower titres, reflecting infants with less than three PCV13 doses and likely detection of maternal antibodies. This is followed by a vaccine-induced increase (positive slope) in IgG until a peak titre, a period of waning (negative slope) until a nadir titre, and a subsequent increase in titre which we hypothesise is due to accumulating natural exposure to *S. pneumoniae*. Serotype 3 showed no population-level vaccine-induced increase and, unique to serotype 3, no post-nadir increase (slope=0), despite evidence of natural exposure (supplementary table 2). As expected there was no vaccine-induced increase in non-VT 33F IgG titres, though a post-nadir increase was observed, suggesting natural exposure. No model could be fitted for serotype 12F because too many measurements were either below the assay’s LLD (73·2%) or had no detectable IgG (16·9%). Three serotypes were associated with a two-stage population-level waning after the post-vaccine peak, with steeper initial waning and later nadirs than other serotypes (including reaching nadir at 26·1 months [9V], 27·0 months [4], and 27.3 months [7F] of age). Serotype 1 included a prolonged post-nadir trough (period of no change in population-level titre [slope=0]) between the ages of 14·1 and 39·5 months before an increase in titre was detected, presumably due to natural exposure in older children.

**Figure 1.**
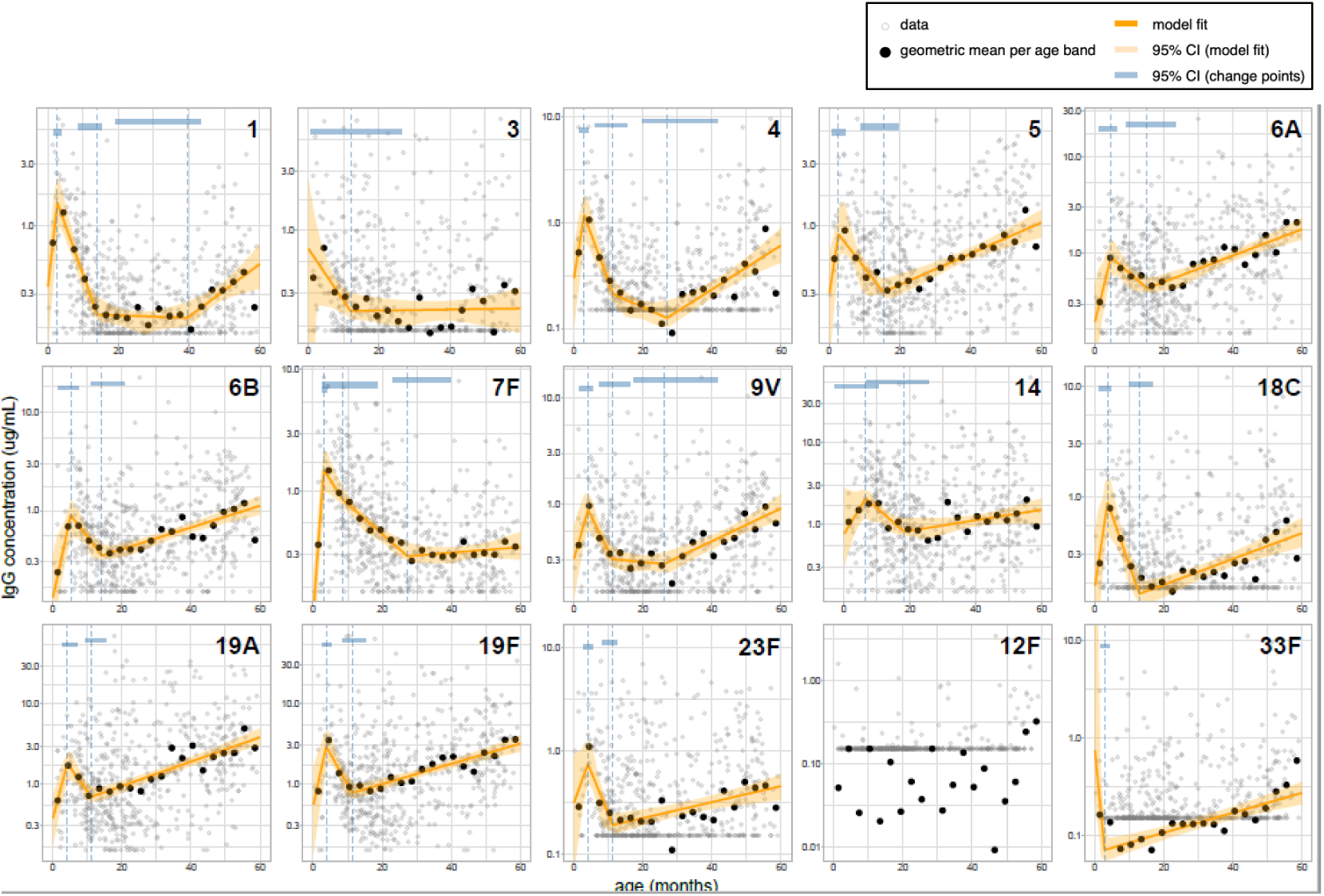
Linear spline regression model evaluating the relationship between age and IgG Grey dots indicate empirical IgG titre datapoints for each sample. Black dots are the geometric mean concentrations (GMCs) for all datapoints within each 3-month age band. The orange line is the spline model fit and the orange shaded area is the 95% CI for the model fit. Vertical dashed lines indicate changepoints. Blue bars at the top of each image are the 95% CI for the age at each changepoint. Y-axes differ in scale, including IgG concentrations from 0–1μg/mL (12F), 0–3μg/mL (serotypes 1, 3, 5), 0–10μg/mL (serotypes 4, 6B, 7F, 9V, 18C, 23F, 33F); 0-30μg/mL (6A, 14, 19A, 19F). The row of grey dots at the bottom edge of each image are datapoints with titres <LLD. This analysis took the actual LLD into account and treated all values ≤0·15μg/mL as censored. Y-axis slope parameters are on the log-IgG scale. Supplementary tables 4A & 4B report the value for each calculated GMC and 95% CI.

**Table 2.** Serotype specific parameter estimate from a population-level spline model analysis

| Sero-<br>type | Profile<sup>1</sup> | Increase in IgG<sup>2</sup><br>Vaccine induced |  |  | Waning IgG<sup>3</sup> |  |  | Increase in IgG,<br>Natural exposure |  |
| --- | --- | --- | --- | --- | --- | --- | --- | --- | --- |
|  |  | Slope to peak<br>95% CI | Age (m) at peak<br>95% CI | [IgG] at peak<br>95% CI | Slope to nadir<sup>4</sup><br>95% CI | Age (m) at nadir<br>95% CI | [IgG] at nadir<br>95% CI | Slope<br>95% CI | [IgG] at final<br>95%CI |
| 1 | 2 | 0.55<br>0.50–0.82 | 2.80<br>1.86–3.81 | 1.5<br>0.75–2.25 | -0.17<br>-0.30–0.14 | 14.10<br>8.82–15.14 | 0.20<br>0.17–0.26 | 0.05<br>0.02–0.07 | 0.51<br>0.33–0.72 |
| 3 | 1 | -0.10<br>-0.40–10.76 | -- | -- | 0.00<br>-0.01–0.02 | 12.00<br>1.03–26.30 | 0.22<br>0.19–0.31 | -- | 0.23<br>0.14–0.34 |
| 4 | 3 | 0.46<br>0.38–0.72 | 3.07<br>1.59–4.27 | 1.18<br>0.55–1.77 | S2: -0.21 (-0.34–0.13)<br>S3: -0.03 (-0.09–0.02) | 27.02<br>19.83–41.36 | 0.13<br>0.10–0.18 | 0.05<br>0.03–0.07 | 0.61<br>0.42–0.90 |
| 5 | 4 | 0.41<br>0.14–0.96 | 2.69<br>1.08–4.43 | 0.86<br>0.51–1.36 | -0.08<br>-0.27–0.05 | 15.34<br>9.11–19.76 | 0.31<br>0.29–0.40 | 0.03<br>0.02–0.04 | 1.07<br>0.82–1.34 |
| 6A | 4 | 0.33<br>0.20–0.63 | 4.71<br>1.37–6.48 | 0.91<br>0.55–1.27 | -0.07<br>-0.24–0.02 | 15.25<br>9.11–23.30 | 0.44<br>0.39–0.58 | 0.03<br>0.02–0.04 | 1.78<br>1.39–2.34 |
| 6B | 4 | 0.38<br>0.19–0.59 | 5.24<br>1.45–7.57 | 0.90<br>0.47–1.25 | -0.11<br>-0.27–0.03 | 14.25<br>11.11–20.94 | 0.35<br>0.32–0.47 | 0.03<br>0.02–0.04 | 1.12<br>0.91–1.41 |
| 7F | 3 | 0.88<br>0.72–1.06 | 3.12<br>2.62–3.90 | 1.47<br>0.82–2.13 | S2: -0.10 (-0.23–0.04)<br>S3: -0.06 (-0.07–0.02) | 27.33<br>23.21–39.78 | 0.29<br>0.27–0.38 | 0.01<br>-0.01–0.03 | 0.35<br>0.27–0.46 |
| 9V | 3 | 0.26<br>0.19–0.48 | 4.19<br>1.77–5.33 | 0.88<br>0.56–1.23 | S2: -0.15 (-0.31–0.07)<br>S3: -0.01 (-0.05–0.03) | 26.14<br>17.39–41.44 | 0.29<br>0.23–0.35 | 0.04<br>0.02–0.06 | 0.91<br>0.70–1.24 |
| 14 | 4 | 0.15<br>-0.01–0.39 | 6.64<br>2.86–10.30 | 2.07<br>1.16–2.63 | -0.08<br>-0.22–0.01 | 18.35<br>6.93–25.79 | 0.82<br>0.72–1.21 | 0.01<br>0.00–0.03 | 1.51<br>1.02–2.10 |
| 18C | 4 | 0.47<br>0.12–1.14 | 3.78<br>1.45–4.76 | 0.87<br>0.43–1.42 | -0.21<br>-0.36–0.10 | 12.95<br>9.89–16.67 | 0.13<br>0.11–0.21 | 0.03<br>0.02–0.04 | 0.47<br>0.35–0.64 |
| 19A | 4 | 0.38<br>0.17–0.57 | 4.22<br>2.79–6.90 | 1.81<br>1.01–2.46 | -0.14<br>-0.34–0.05 | 11.24<br>9.47–15.48 | 0.70<br>0.60–0.97 | 0.04<br>0.03–0.04 | 3.86<br>3.06–4.96 |
| 19F | 4 | 0.45<br>0.16–0.80 | 3.79<br>2.58–5.39 | 2.88<br>1.38–4.38 | -0.17<br>-0.42–0.10 | 11.34<br>8.61–15.12 | 0.78<br>0.67–1.10 | 0.03<br>0.02–0.04 | 3.17<br>2.51–4.01 |
| 23F | 4 | 0.21<br>-0.02–0.67 | 4.17<br>2.96–5.59 | 0.74<br>0.46–1.25 | -0.19<br>-0.53–0.15 | 11.34<br>8.22–12.46 | 0.19<br>0.16–0.25 | 0.02<br>0.01–0.03 | 0.45<br>0.33–0.58 |
| 12F* | 1 | -- | -- | -- | -- | -- | -- | -- | 0.27<br>0.21–0.35 |
| 33F | 1 | -- | - No Peak- | - No Peak- | -- | 2.90<br>1.76–4.17 | 0.07<br>0.06, 0.14 | -- | -- |

<sup>1</sup>Profiles are visually defined (ie, not based on criteria of statistical significance) to help the reader categorise trends: 1=fitted model estimated no vaccine-induced IgG increase, 2=fitted model estimated a trough post nadir for Serotype 1, indicating no change in IgG titre from 14.0 to 39.5 months of age, 3=fitted model estimated two unique slopes within the waning phase, 4=fitted model estimated two changepoints—one at IgG peak and one at nadir prior to an increase in titre presumably due to natural exposure; \*No model could
be fitted for serotype 12F as almost all measurements were below the detection limit. <sup>2</sup>Though increase in IgG can include IgG induced by natural exposure, we hypothesise that this stage is driven largely by vaccine-induced IgG. <sup>3</sup>The aggregate dynamic of waning IgG will be influenced by waning of maternal and vaccine-induced IgG and increased IgG induced by natural exposure. <sup>4</sup>Nadir refers to the minimum population-level IgG titre estimated. ND=not defined by the model. [IgG] = concentration of serotype-specific IgG is µg/mL. S2 and S3: the second (S2) and third (S3) slopes estimated by the linear spline model, in these cases both (S2 and S3) occur during post-vaccination waning.

As shown in table 2, the age range at peak IgG (after vaccination) was from 2·8 months (serotype 1) to 6·6 months (serotype 14). The age range at nadir (after vaccine-induced peak) was from 11·2 months (19F and 23F) to 27·3 months (7F). The IgG titre at nadir ranged from 0·13μg/mL (4 and 18C) to 0·82μg/mL (14).

### Seroincident events

An analysis of the 82 pairs of linked samples, censored to include only children at least 6 months of age, showed evidence of seroincident events (ie, higher IgG titre at secondary sample relative to primary sample) during follow-up (supplementary table 5 and supplementary figure 1). Serotype 5, for example, had the highest proportion of seroincident events, with 48·6% (36/74) of linked sample pairs indicating an increase in IgG titre: 10·8% (n=8) with a ≥2-fold increase and 4·0% (n=3) with a ≥4-fold increase. Evidence of increasing incidence of exposure events with age was further supported by NanA and Ply protein-specific antibody profiles (supplementary figure 2A-2C). Protein responses start increasing at roughly the same age as post-nadir anti-capsular IgG, with similar trends reported by Turner and colleagues in Thailand.^24^

### Proportion of samples attaining CoPs

Figure 2 shows the proportions of samples attaining CoPs associated with prevention of IPD and carriage. The data used to develop this figure are reported in supplementary table 7. CoPs associated with preventing disease include 0·35μg/mL and serotype-specific CoPs reported by Andrews and colleagues.^8^ CoPs associated with preventing carriage include serotype-specific CoPs reported by Voysey and colleagues.^12^ Except for serotypes 7F (lowest rate of increase [slope=0·1] in post-nadir IgG concentration) and serotype-3, the proportion of children attaining the aggregate 0·35μg/mL CoP against IPD, for example, dropped until around the 1-year age group, increasing thereafter through the 4-year age group. A similar population trend was demonstrated for all CoP thresholds evaluated.

**Figure 2.**
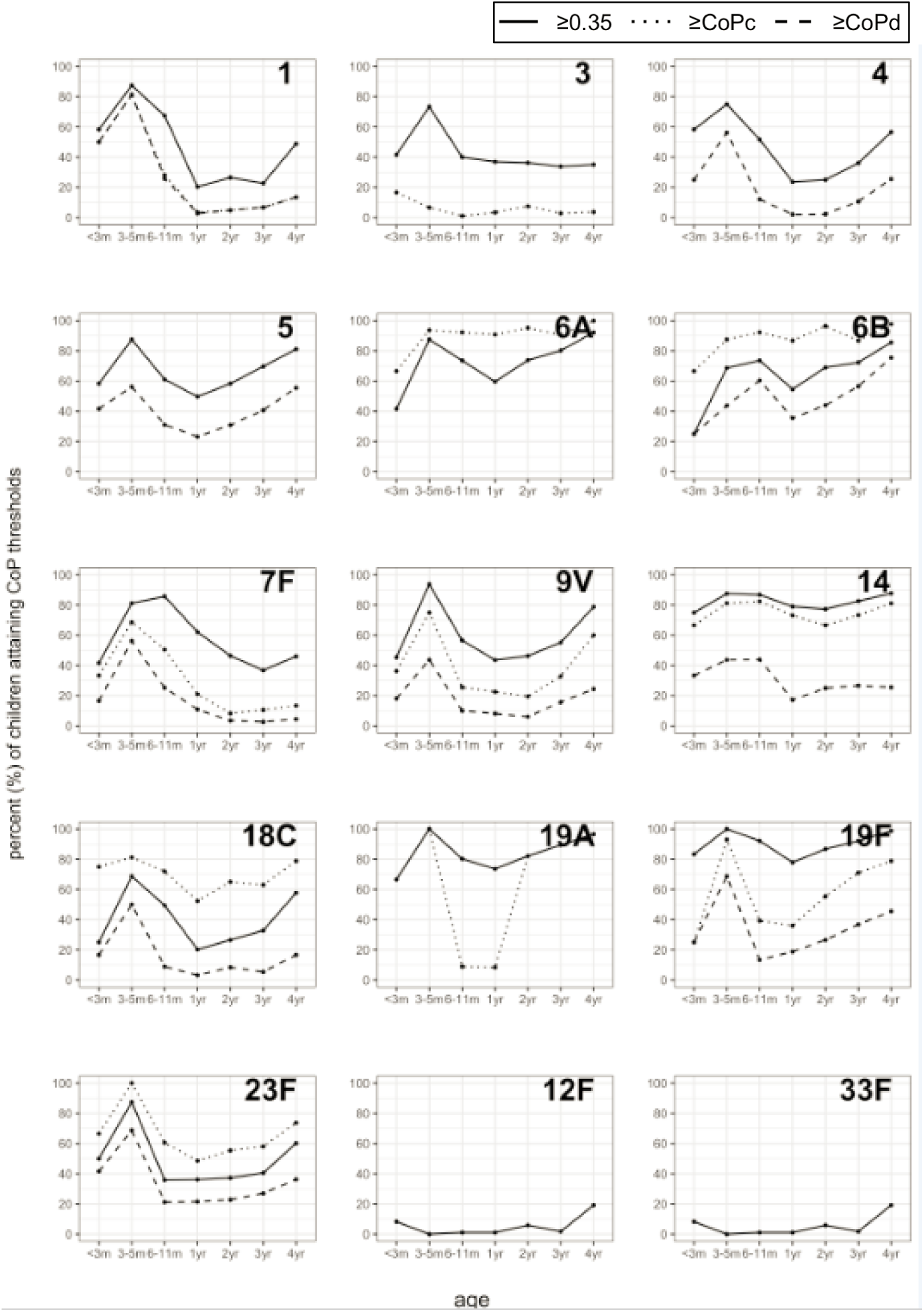
Percent of children attaining CoP thresholds (μg/mL) after PCV13 vaccination, stratified by age group Solid lines indicate percent of children in with a serotype-specific IgG level exceeding 0·35μg/mL. Dashed lines indicate percent of children with a serotype-specific IgG level exceeding the putative IPD CoP reported by Andrews and colleagues: serotype 1=0·78, 3=2·83, 4=0·35, 5=undefined, 6A=0·16, 6B=0·16, 7F=0·87, 9V=0·62, 14=0·46, 18C=0·14, 19A=1·00, 19F=1·17, 23F=0·20; Dotted lines indicate percent of children with a serotype-specific IgG level exceeding the putative carriage correlate of protection reported by Voysey and colleagues: serotype 1=0·81, 3=undefined, 4=1·16, 5=0·73; 6A=undefined, 6B=0·50, 7F=1·60, 9V=1·31, 14=2·48, 18C=1·32, 19A=undefined, 19F=2·54, 23F=0·63. Note that some 15 serotypes (1, 3, 4, 5, 6A, 19A, 12F, 33F) have no putative CoP data, reflected by having fewer lines. m=months, yr=years. Refer to supplementary table 7 for the values used to develop this figure.

### Regression model to define population-level correlates of protection against carriage

Based on community carriage prevalence after PCV13 introduction, reported by the carriage^15^ (supplementary table 2), VTs were categorised as “lower transmission” if carriage prevalence was <1% (4, 6B, 7F, 9V, 18C) and “higher transmission” if ≥1% (6A, 14, 19A, 19F, 23F). We excluded serotypes 1 and 5 (high IPD prevalence despite low carriage prevalence in this setting) and serotype 3 (PCV13 reportedly confers less protection than for other VT).^27^

For lower-transmission VTs, all except 9V were associated with no or low carriage among the 3 to 6 month and 6 to 12 month age groups. This indicates interrupted transmission in these age groups at the indicated serotype-specific median titres (supplementary table 6), indicating the approximate carriage CoP is below 1·2μg/mL. Figure 3A further refines this approximation, indicating that the linear regression estimates flat or decreased relationships between carriage and median serological titres across all age groups, with a constant 0·50–0·60μg/mL estimated as the carriage CoP. This is consistent with the findings of Voysey and colleagues for these serotypes (0·50–1·60μg/mL).

**Figure 3.**
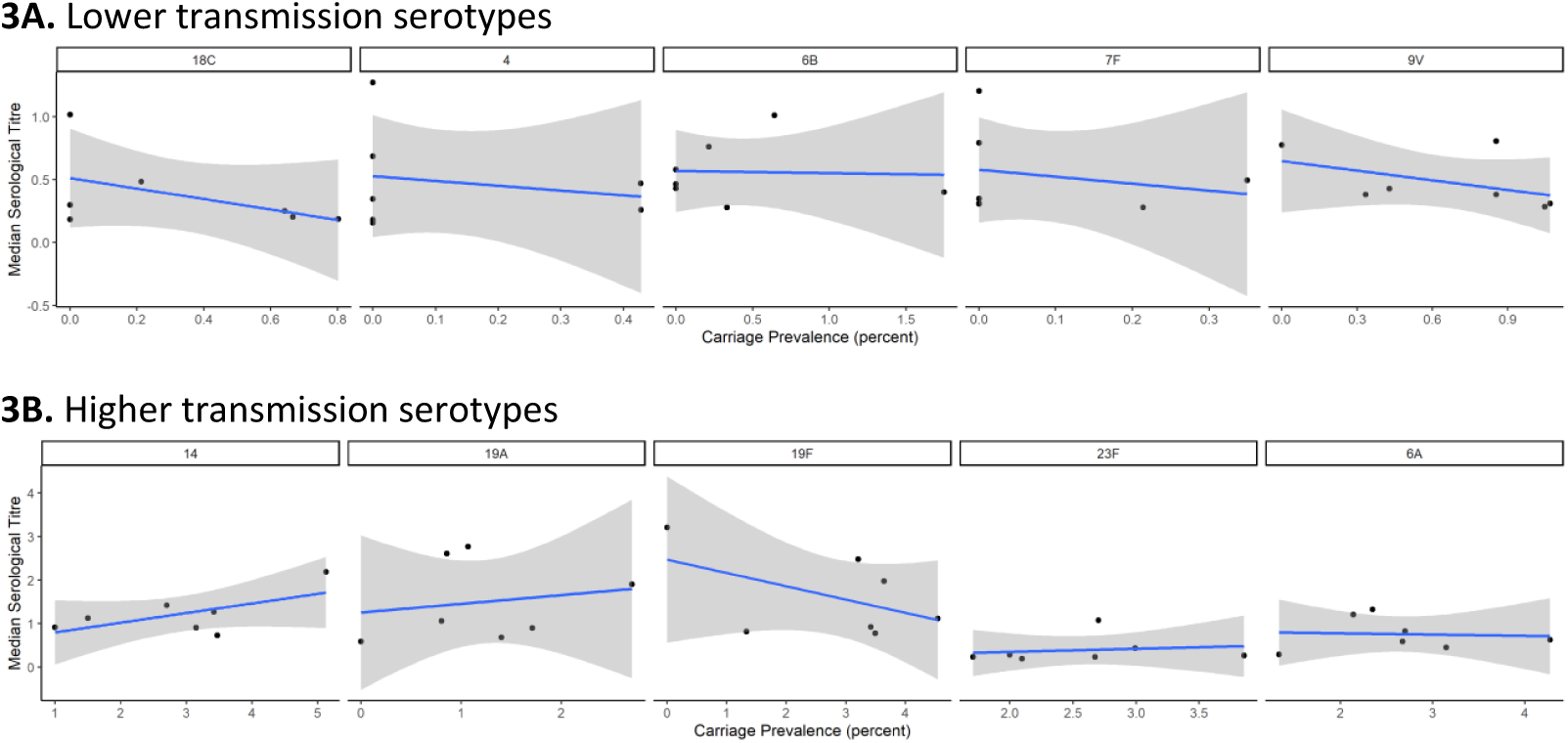
Median serological titre and carriage prevalence for children under 5 years old To infer CoP, we used a linear regression between median serotype-specific IgG titre (y-axis) and carriage prevalence (xaxis), defining the CoP as the value of the fitted regression line at 0% prevalence. Blue lines indicate linear regression lines. Grey shaded area indicates the standard error bounds.

For higher transmission VTs, there was intermittent transmission in the 3 to 6 months age group, indicating an approximate CoP of 1·0μg/mL for serotypes 6A and 23F, 1·5μg/mL for 14, 2·0μg/mL for 19A, and 3·0μg/mL for 19F. The linear regression model (figure 3B) was unable to estimate a clear association between carriage and median serological titre, likely due to ongoing high transmission and natural boosting. However, in reviewing serotype-specific titre trends associated with reduced carriage (Figure 3B), we estimated IgG titres associated with reduced carriage, including 1·0μg/mL for 6A and 23F (as reported above), 2·5μg/mL for 14 and 19F, and 2·0μg/mL for 19A (supplementary Table 1).

### Age period of potential vulnerability to pneumococcal carriage and IPD

By aligning our serotype-specific immunogenic profiles with putative CoPs, we estimated population-level titres that drop below IPD and carriage CoPs for extended periods before cumulative natural exposure induces an increase in titres (table 4). Population-level titres of nine VTs fell below 0·35μg/mL for a range of 0·2– 51·9 months (median 22·9). The titres of nine VTs fell below the serotype-specific CoP for IPD for a range of 5·0–52·0 months (median 24·8). For available carriage CoPs, titres fell below the serotype-specific CoP for all 10 VTs reported by Voysey and colleagues (range: 17·6–54·0 months, median 55·4) and also for CoPs estimated from this study’s empirical data (range: 24·0–51·0 months, median 40·8). Supplementary figure 3 shows the spline model output for 23F, as an example, with ages below the CoPs indicated.

**Table 4.** Estimated age range (months) after vaccine-induced peak when IgG titres are below the IPD and carriage CoPs among children

|  | IPD |  | Carriage |  |
| --- | --- | --- | --- | --- |
|  | Age (mo) range below 0.35µg/mL | Age (mo) range below serotype-specific CoP-d<sup>¶</sup> | Age (mo) range below serotype-specific CoP-c<sup>‡</sup> | Age (mo) range below serotype-specific CoP-c<sup>¥</sup> |
| 1 | 11.1 – 52.0 | 7.0 – 59+ | 6.3 – 59+ | UND |
| 3 | 7.1 – 59+<sup>a</sup> | All ages <60<sup>a</sup> | UND | UND |
| 4 | 9.0 – 48.4 | 10.5 – 44.0 | 3.0 – 59+ | 6.5 – 58.0 |
| 5 | 13.9 – 19.7 | UND | 4.8 – 46.3 | UND |
| 6A | No ages <60 | No ages <60 | UND | 8.0 – 32.0 |
| 6B | 14.2 – 14.4 | No ages <60 | 10.8 – 28.4 | 9.5 – 34.0 |
| 7F | 24.2 – 59+ | 9.5 – 59+ | All ages <60 | 18.5 – 59+ |
| 9V | 10.3 – 33.2 | 6.0 – 48.5 | All ages <60 | 8.0 – 43.0 |
| 14 | No ages <59 | No ages <60 | All ages <60 | All ages <60 |
| 18C | 8.2 – 49.3 | 12.0 – 22.5 | All ages <60 | 7 – 59+ |
| 19A | No ages <60 | 7.5 – 21.5 | UND | All ages <41.0 |
| 19F | No ages <60 | 10.0 – 26.0 | 4.6 – 59+ | 4.0 – 54.0 |
| 23F | 8.1 – 45.8 | 11.0 – 16.0 | 5.0 – 59+ | All ages <60 |

<sup>¶</sup>Putative CoP for IPD (Andrews et al): serotype 1=0.78µg/mL, 3=2.83, 4=0.35, 5=undefined, 6A=0.16, 6B=0.16, 7F=0.87, 9V=0.62, 14=0.46, 18C=0.14, 19A=1.00, 19F=1.17, 23F=0.20. <sup>‡</sup>Putative CoP for carriage (Voysey and colleagues.) limited to PCV10 serotypes: serotype 1=0.81, 3=undefined, 4=1.16, 5=0.73; 6A=undefined, 6B=0.50, 7F=1.60, 9V=1.31, 14=2.48, 18C=1.32, 19A=undefined, 19F=2.54, 23F=0.63. <sup>¥</sup>Putative CoP for carriage, based on empirical study data: serotype 1=undefined, 3=undefined, 4=0.5, 5=undefined; 6A=0.8, 6B=0.50, 7F=0.50, 9V=0.50, 14=2.0, 18C=0.5, 19A=2.0, 19F=3.0, 23F=1.0. <sup>a</sup>Model estimates no vaccine-induced increase in anti-serotype 3 antibody. CoP-c=correlate of protection for carriage, CoP-d=correlate of protection for disease, IPD=invasive pneumococcal disease, mo=month, UND=Undefined.

## DISCUSSION

In this under-5-year-old population in Malawi, a 3+0 PCV13 vaccine schedule with a catch-up-campaign among children aged under-1-year, vaccine-induced antibody against multiple VTs was not sustained at a population level beyond the first year of life. Despite a post-nadir increase, presumably due to natural exposure, the antibody concentrations of several VTs remained below putative CoPs for both IPD and carriage for extended periods. This likely contributes to the previously reported residual circulation of VT and VT-IPD.^15,18^ Indeed, the absence of population immunity against serotype 3 (a commonly carried VT) and the extended period during which immunity against serotype 1 (uncommonly carried but a prominent cause of disease) falls below the CoPs for IPD and carriage after the first year of life are of particular concern.

This population-level analysis of age-associated anti-capsular immunity reveals antibody profiles that are largely consistent across VTs except for serotype 3. There is a vaccine-induced increase in IgG that peaks at around 5 (range 2.7–6.6) months of age for the 12 serotypes which, according to several efficacy trials and post-introduction effectiveness studies, is likely to be protective.^17,28^ However, this peak is not sustained, falling to a nadir at around 14 (range 11.2–27.3) months of age for the 12 serotypes. Our hypothesis that the post-nadir increase in titres is due to accumulating natural exposure is supported by evidence of persistent carriage of all 13 VT and consequent sero-incident events for commonly carried serotypes. The heterogeneity between individual VTs in the rates of vaccine-induced IgG production and waning in relation to the control of pneumococcal colonisation has received insufficient attention to date.

Serotype-specific immunogenic profiles among infants are complex, comprising a combination of waning maternal antibody and antibodies induced by both vaccine and natural exposure. Previous studies have shown varying prevalences of maternal pneumococcal antibodies, for example, highest for serotypes 14 and 19F (92% and 80%, respectively) and lowest for serotypes 4 and 1 (30% and 34%, respectively).^29^

For serotype 3, we saw no evidence of a population-level vaccine-induced antibody and, despite a relatively high prevalence of carriage, unique to serotype 3, no post-nadir increase up to 59 months of age. This was consistent with previous evidence that PCV13 confers less and shorter-term protection against serotype 3 among vaccinated children relative to other VTs.^27^

In our evaluation using empirical study data, we report similar CoP thresholds for carriage to those reported by Voysey and colleagues.^12^ Voysey and colleagues also reported protective antibody concentrations against carriage being, on average, twice as high in LMICs than in HICs.

Recognizing that the aggregate 0·35μg/mL CoP for IPD was derived from pooled infant data limited to high-income (USA) and upper middle-income (South Africa) country settings, regional differences should be considered when evaluating CoPs for carriage and disease,^7^ especially in settings such as Malawi where VTs continue to contribute to pneumococcal carriage and IPD disease burden. Crucially, our data highlight the potential loss of the protective benefit of a 3+0 schedule by the second year of life which, apart from capsule-independent immunity, presents a potential window of vulnerability to carriage and disease.

While this novel population-based analysis was both cost-effective and less complex than a longitudinal cohort study, there were limitations, including the extrapolation of findings to the individual level. While we are not aware of a similar population-based study to which we can compare our findings, Turner and colleagues presented a similar analysis with similar serological profiles from their work in Thailand.^24^ However, their study was completed before PCV implementation and was limited to children <2 years of age. Additionally, the interpretation of CoP for pneumococcal carriage remains associated with considerable uncertainty. Further analysis of carriage and serological data from other similar geospatial and temporal settings will strengthen these estimates.

In conclusion, in the context of a 3+0 PCV13 schedule, we report the absence of a sustained vaccine-induced antibody response to multiple VTs beyond the first year of life. This may result in a window of vulnerability to both pneumococcal carriage and disease. With evidence of residual VT-IPD in settings such as Malawi, including among highly vulnerable neonates,^30^ strategies to counter this residual burden are required to achieve sustained vaccine-induced antibody titres and, therefore, control. An extended catch-up campaign at the time of PCV introduction can speed-up the reduction in VT carriage. A booster dose, also endorsed by the WHO, given later in the first year or second year of life may also be used to overcome these factors. However, both the impact and the cost-effectiveness of such strategies need to be evaluated before they are implemented in vaccine programmes.

## Data Availability

Deidentified individual participant data and a data dictionary can be made available on request to the corresponding author with a research proposal and signed data usage agreement.

https://github.com/gitMarcH/PCVPA_IgGCensoredRegressionSplineModelling

## Contributors

TDS, NF, and RSH conceived the study. TDS, NF, and RSH designed the study, with contributions from JEM and MG. TDS, JM, MM, CB, EP, and DG oversaw laboratory activities. TDS, JEM, DT, BM, and AAM oversaw data curation. TDS, MYRH, and SB completed formal analysis. TDS, JEM, MYRH, and RSH had full access to all the data in the study. TDS and RSH jointly wrote the first draft of the manuscript. All authors have contributed to data interpretation while editing and commenting on the draft manuscript. All authors have read and approved the final manuscript. TDS, together with the NF and RSH, had final responsibility for the decision to submit for publication.

## Data sharing

Deidentified data can be made available on request to the corresponding author with a research proposal and signed data usage agreement.

## Acknowledgments

This work was funded by Bill & Melinda Gates Foundation (OPP1117653 to RSH), a Wellcome Programme Grant (WT091909/B/10/Z to RSH), and National Institute for Health & Care Research (NIHR) Global Health Research Unit on Mucosal Pathogens using UK aid from the UK Government (16/136/46 to RSH). The Malawi–Liverpool–Wellcome Programme is supported by a Strategic Award from Wellcome, U.K. (206545/Z/17/Z). TDS, RSH and NF are supported by the NIHR Global Health Research Unit on Mucosal Pathogens using UK aid from the UK Government. The findings and the views expressed are those of the authors and not necessarily those of the NIHR or any other affiliated institutions. We acknowledge Dr Hao Hu for his dedicated work in leading the development of the regression model used to report correlates of protection for carriage using the study’s empirical data.

