## Supplementary material for "Waning of PCV13 vaccine-induced antibody levels within the first year of life, using a 3+0 schedule: an observational population-level serosurveillance study among children under 5 years old in Blantyre, Malawi"

**Supplementary Material**
**Swarthout TD and colleagues**

**Title:** Waning of PCV13 vaccine-induced antibody levels within the first year of life, using a 3+0 schedule: an observational population-level serosurveillance study among children under 5 years old in Blantyre, Malawi.

### Table of Contents

Table S1 - Correlates of protection (µg/mL) for invasive pneumococcal disease and carriage ……………………….…………………………. 3
Table S2 - Community carriage serotype distribution among children aged <5 years, PCVPA Study ……………………….……..................... 4
Table S3 - Assay output summary ………………………………………………………………………………………………………. 5
Table S4 - Serotype-specific GMCs calculated using empirical data as shown in manuscript Figure 1, stratified by 3-month age groups …….. 8-9
Table S5 - Trend among paired (primary and secondary) samples, censored for ages <6 months ………………………………………....... 10
Table S6 - Median IgG titre and carriage prevalence (rounds 4–6 in PCVPA Study) in the 3 to 6 months and 6 to 12 months age groups ……... 13
Table S7 - Table S7. Percent of children attaining CoP thresholds (µg/mL) post vaccination, stratified by age group ……………………..... 14
Figure S1 - Seroincident events, based on 82 linked pairs ………………………………………………………………………................. 11
Figure S2 - Protein-specific antibody profiles, stratified by 3-month age groups ………………………………………………………………… 12
Figure S3 - Population anti-23F titre trends, age periods when above or below CoP ……………………………………………………………….. 15
Supplementary methods, linear spline regression model ………………………………………………………………………………………… 6-7
References …………………………………………………………………………………………………………………………….. 16

**Table S1 -** Correlates of protection (μg/mL) for invasive pneumococcal disease and carriage

| | IPD | | Carriage | |
|---|---|---|---|---|
| | **Siber[1] μg/mL** | **Andrews[2] μg/mL 95%CI** | **Voysey[3] μg/mL 95%CI** | **Empirical, Malawi study μg/mL** |
| 1 | 0·35 | 0·78<br>0·47–1·68 | 0·81<br>0·54–1·22 | |
| 3 | 0·35 | 2·83<br>1·16–∞ | | |
| 4 | 0·35 | 0·35<br>0·20–1·17 | 1·16<br>0·90–1·49 | 0·50-0·60 |
| 5 | 0·35 | ND | 0·73<br>0·42–1·26 | |
| 6A | 0·35 | 0·16<br>0·08–1·05 † | | 1·0 |
| 6B | 0·35 | 0·16<br>0·08–2·54 | 0·50<br>0·40–0·61 | 0·50-0·60 |
| 7F | 0·35 | 0·87<br>0·40–1·80 | 1·60<br>1·25–2·05 | 0·50-0·60 |
| 9V | 0·35 | 0·62<br>0·19–∞ | 1·31<br>1·08–1·58 | 0·50-0·60 |
| 14 | 0·35 | 0·46<br>0·25–1·12 | 2·48<br>2·02–3·04 | 2·5 |
| 18C | 0·35 | 0·14<br>0·09–0·40 | 1·32<br>0·95–1·82 | 0·50-0·60 |
| 19A | 0·35 | 1·00<br>0·60–2·47 | | 2·0 |
| 19F | 0·35 | 1·17<br>0·62–4·62 | 2·54<br>1·83–3·53 | 2·5 |
| 23F | 0·35 | 0·20<br>0·08–1·50 | 0·63<br>0·47–0·83 | 1·0 |
| 12F | | | | |
| 33F | | | | |

Shaded grey cells represent serotypes that were not evaluated in the respective analyses.

**Table S2 -** Community carriage serotype distribution among children aged <5 years, PCVPA Study

| Serotype | n | Carriage prevalence | Contribution to total VT carriage n=719 |
|---|---|---|---|
| VT§ (n=4 234) | | | |
| 1 | 13 | 0·31 | 1·81 |
| 3 | 122 | 0·88 | 16·97 |
| 4 | 12 | 0·28 | 1·67 |
| 5 | 7 | 0·17 | 0·97 |
| 6A | 89 | 2·10 | 12·38 |
| 6B | 27 | 0·64 | 3·76 |
| 7F | 3 | 0·07 | 0·42 |
| 9V | 29 | 0·68 | 4·03 |
| 14 | 91 | 2·15 | 12·66 |
| 18C | 16 | 0·38 | 2·23 |
| 19A | 44 | 1·04 | 6·12 |
| 19F | 146 | 3·45 | 20·31 |
| 23F | 120 | 2·83 | 16·69 |
| NVT‡ (n=1 629) | | | |
| 12F | 16 | 0·98 | ND |
| 33F | 12 | 0·73 | ND |

§Source: Pneumococcal carriage surveillance, 2015–2019, latex agglutination (partly reported in Swarthout TD and colleagues[4]). ‡Source: Pneumococcal carriage surveillance, 2015–2017, microarray (partly reported in Swarthout TD and colleagues[5]). <5 years: includes PCV age-ineligible (pre-vaccine, 4–8 weeks old) and PCV13-vaccinated (3–4 years old). ND=Not done, NVT=non-vaccine serotype, VT= vaccine serotype.

**Table S3 -** Assay output summary

| Serotype | < LLD[¶] n (%) | NR[§] N (%) | QNS[‡] n (%) | Samples in analyses n (%) | In primary analysis, n[!] | In secondary analysis, n pairs | Maximum titre (median)[#] μg/mL |
|---|---|---|---|---|---|---|---|
| 1 | 132 (20·7) | 22 (3·5) | 1 (0·2) | 615 (96·4) | 534 | 72 | 6·63 (0·25) |
| 3 | 202 (31·7) | 36 (5·6) | 3 (0·5) | 599 (93·9) | 519 | 70 | 8·04 (0·24) |
| 4 | 217 (34·0) | 0 (0·0) | 2 (0·3) | 636 (99·7) | 554 | 76 | 9·64 (0·24) |
| 5 | 52 (8·2) | 3 (0·5) | 1 (0·2) | 634 (99·4) | 553 | 74 | 6·72 (0·47) |
| 6A | 43 (6·7) | 0 (0·0) | 1 (0·2) | 637 (99·8) | 555 | 76 | 24·55 (0·66) |
| 6B | 62 (9·7) | 3 (0·5) | 1 (0·2) | 634 (99·4) | 552 | 76 | 22·66 (0·50) |
| 7F | 55 (8·6) | 0 (0·0) | 3 (0·5) | 635 (99·5) | 553 | 75 | 8·66 (0·43) |
| 9V | 112 (17·6) | 9 (1·4) | 2 (0·3) | 627 (98·3) | 546 | 72 | 15·49 (0·40) |
| 14 | 30 (4·7) | 2 (0·31) | 1 (0·2) | 635 (99·5) | 554 | 75 | 63·49 (1·13) |
| 18C | 219 (34·3) | 11 (1·7) | 2 (0·3) | 625 (98·0) | 546 | 71 | 11·92 (0·24) |
| 19A | 19 (3·0) | 0 (0·0) | 1 (0·2) | 637 (99·8) | 555 | 76 | 71·31 (1·19) |
| 19F | 9 (1·4) | 4 (0·6) | 2 (0·3) | 632 (99·1) | 552 | 74 | 69·28 (1·29) |
| 23F | 204 (32·0) | 7 (1·10) | 3 (0·5) | 628 (98·4) | 547 | 73 | 12·90 (0·27) |
| 12F | 467 (73·2) | 108 (16·9) | 2 (0·3) | 528 (82·8) | 455 | 64 | 3·37 (LLD) |
| 33F | 348 (54·6) | 7 (1·10) | 1 (0·2) | 630 (98·7) | 549 | 75 | 11·02 (LLD) |
| PsaA | 0 | 0 | 1 | 637 (98·7) | 555 | - | 13846·96 |
| NanA | 0 | 0 | 1 | 637 (98·7) | 554 | - | 8842·46 |
| Ply | 0 | 1 | 1 | 636 (98·7) | 555 | - | 3873·81 |

[¶]<LLD (less than the lower limit of detection): refers to a value below the assay's lower limit of detection. Samuples <LLD do have detectable IgG, but the ELISA assay for IgG quantification cannot accurately determine the concentration value. The LLD was <0·150 μg/mL. [§]NR (no result): ELISA sample results that were non-parallel (NP) were then retested. If the retest was also non-parallel, then a result of NR (no result) was recorded. [‡]QNS (quantity not sufficient): There was inadequate volume of sample to measure all targeted antibodies. [!]Total in analyses=638-(NR+QNS). [#]Maximm and median IgG for primary plasma samples only.

**Supplementary methods, linear spline regression models:** A flexible modelling framework using linear splines for censored regression models to estimate serological profiles

**Lower limit of detection; censored regression:** The assay's lower limit of detection (LLD, 0·15 µg/mL) means that observations below the detection limit (DL) are left-censored. To account for this, we use censored regression. Specifically, we assume a Gaussian linear model for $y$, the natural logarithm of the measured IgG concentration, with a single explanatory variable, $x$, age:

$$y = \beta_0 + \beta_1 x + \epsilon = f(x, \boldsymbol{\beta}) + \epsilon \qquad (1)$$

where $\epsilon \sim \mathcal{N}(0, \sigma^2)$.

We do not directly observe $y$, but only $y_M$, where values below the detection limit are censored:

$$y_M = \begin{cases} y, & y > \tau \\ \tau, & y \leq \tau \end{cases}$$

where $\tau$ is the lower limit of detection ($\tau = 0.15$µg/mL in our data).

Since we assume a Gaussian model, this allows us to write down, for an observed data set $\{x_i, y_{M,i}\}_{i=1}^{n}$, the log-likelihood below[6], thereby making the usual maximum likelihood inference theory available:

$$l(\beta, \sigma^2) = \sum_{y_{M,i} > \tau} \log\left(\phi\left(\left(y_{M,i} - f(x, \boldsymbol{\beta})\right)/\sigma\right)\right) + \sum_{y_{M,i} \leq \tau} \log\left(\Phi(f(x, \boldsymbol{\beta}) - \tau)\right) \qquad (2)$$

**Identify and estimate change points; linear regression splines:** To address the objectives, we needed to fit a model that allows the linear relationship between age and IgG to change over time. A natural candidate for this is linear splines. Splines fit piece-wise linear regression models, allowing the slope to change at specific values of the predictor variable. These locations, commonly referred to as knots, define intervals over which the relationship between predictor and response is linear.

Mathematically, for a response $y$ and a predictor $x$, a linear spline with 3 knots at locations $x = k_1, k_2, k_3$ can be written down as:

$$y = \beta_0 + \beta_1 \cdot x + \beta_2 \cdot (x - k_1)_+ + \beta_3 \cdot (x - k_2)_+ + \beta_4 \cdot (x - k_3)_+ + \epsilon \qquad (3)$$

where $(x)_+ = x$ if $x \geq 0$ and 0 otherwise.

This means that the fitted curve has different slopes in different intervals of values of $x$:

| interval | Slope |
|---|---|
| (-Inf, $k_1$) | $\beta_1$ |
| [$k_1$,$k_2$) | $\beta_1 + \beta_2$ |
| [$k_2$,$k_3$) | $\beta_1 + \beta_2 + \beta_3$ |
| [$k_3$,Inf) | $\beta_1 + \beta_2 + \beta_3 + \beta_4$ |

**Statistical inference for number and locations of knots (bootstrap and AIC):** Usually splines are used to produce smooth functional fits (typically one would use cubic rather than linear functions) and knot locations are either user-provided or set to regular intervals in the predictor variable.

For the purpose of the pneumococcus serology data however, the number and the locations of the knots, i.e., the change points, are of interest themselves and we would like to do statistical inference on the knot locations. To this end, we treat the knot locations $k_1, k_2, k_3, \ldots$ as model parameters and estimate them using numerical optimisation. We then use the bootstrap to derive confidence intervals for the locations of the knots.

To identify which number of change points best fits the data, we fit models with several numbers of knots, then use Akaike Information Criterion (AIC) to select the optimal number of knots.

**Putting it all together:** Given the nature of the data (IgG titre with a lower detection limit) and the fact that we consider several plausible biological models for vaccine response profiles (with changing slopes) for which it is important to know when the changes in IgG levels occur, we need our modelling framework to satisfy several requirements:

1. Properly account for data values that are left censored (below the detection limit).
2. Treat the knot locations as parameter values to be estimated during model fitting.
3. Return a fit for the geometric mean IgG concentration as a function of age.
4. Select the optimal number of knots / changepoints.

This means that our model is (1), with $f(x, \boldsymbol{\beta})$ in (1) replaced with $f(x, \boldsymbol{\beta}, \boldsymbol{k})$ using the functional form from (3).

In summary, the key characteristics of our modelling framework are:

1. We fit censored regression models with linear splines for the age predictor variable.
2. Knot locations are treated as model parameters and fitted by numerical minimisation of a least squares function. Since the model is now no longer fitted by maximum likelihood, we use bootstrap techniques to derive confidence intervals for slope parameters (as well as predicted average concentration at any given age) and knot locations. Specifically, the bootstrap percentile method[7] is used for CIs.
3. Before model fitting, the IgG data are log-transformed, so that the fitted arithmetic mean corresponds to the log of the geometric mean on the original data scale (i.e. we can exponentiate the fitted curve to have a fit for the geometric mean).
4. For every serotype, models are fitted with $k = 0,1,2,3$ knots / changepoints and the optimal value for $k$ is selected using AIC.

Our implementation was done in the R environment for statistical programming, making use of packages rms[8] (for the linear spline terms), censReg[11] (for the censored regression likelihood), boot[7,10] (for the bootstrapped CIs) and custom code to estimate model parameters and selecting the number of knots / changepoint.

**Conclusion:** We present a flexible modelling framework to estimate both the number and location of changepoints in serological profiles. Key benefits of our approach are the principled statistical analysis of serological profile data which are increasingly common and the grouping of profiles, giving insights into the underlying biology.

The modelling code is made available to other researchers on GitHub[11]

**Table S4A -** Serotype-specific GMCs calculated using empirical data as shown in manuscript Figure 1, stratified by 3-month age groups

| **Age band (months)** | n | **Serotype 1** | **Serotype 3** | **Serotype 4** | **Serotype 5** | **Serotype 6A** | **Serotype 6B** | **Serotype 7F** |
|---|---|---|---|---|---|---|---|---|
| 0–02 | 12 | 0·74 (0·30–1·84) | 0·41 (0·13–1·24) | 0·52 (0·22–1·25) | 0·56 (0·30–1·05) | 0·31 (0·15–0·66) | 0·24 (0·11–0·53) | 0·36 (0·15–0·91) |
| 03–05 | 16 | 1·27 (0·74–2·16) | 0·71 (0·42–1·20) | 1·07 (0·55–2·09) | 0·93 (0·60–1·45) | 0·90 (0·55–1·48) | 0·69 (0·36–1·32) | 1·49 (0·80–2·76) |
| 06–08 | 41 | 0·66 (0·52–0·83) | 0·31 (0·25–0·39) | 0·47 (0·36–0·62) | 0·57 (0·43–0·76) | 0·70 (0·50–0·98) | 0·70 (0·53–0·92) | 0·97 (0·74–1·27) |
| 09–11 | 49 | 0·39 (0·32–0·48) | 0·28 (0·20–0·41) | 0·28 (0·22–0·36) | 0·40 (0·33–0·49) | 0·58 (0·46–0·73) | 0·49 (0·38–0·64) | 0·82 (0·65–1·03) |
| 12–14 | 40 | 0·24 (0·20–0·28) | 0·23 (0·15–0·36) | 0·22 (0·17–0·27) | 0·44 (0·36–0·54) | 0·59 (0·46–0·75) | 0·42 (0·34–0·51) | 0·60 (0·46–0·78) |
| 15–17 | 54 | 0·21 (0·17–0·26) | 0·27 (0·18–0·41) | 0·15 (0·10–0·22) | 0·32 (0·25–0·41) | 0·46 (0·36–0·59) | 0·37 (0·30–0·45) | 0·48 (0·40–0·57) |
| 18–20 | 43 | 0·20 (0·15–0·26) | 0·20 (0·15–0·27) | 0·17 (0·13–0·23) | 0·35 (0·28–0·45) | 0·51 (0·38–0·68) | 0·40 (0·30–0·53) | 0·48 (0·35–0·66) |
| 21–23 | 47 | 0·19 (0·16–0·24) | 0·22 (0·15–0·33) | 0·15 (0·10–0·22) | 0·38 (0·29–0·50) | 0·44 (0·31–0·63) | 0·40 (0·29–0·56) | 0·40 (0·30–0·52) |
| 24–26 | 26 | 0·24 (0·16–0·34) | 0·18 (0·09–0·34) | 0·11 (0·05–0·23) | 0·33 (0·24–0·45) | 0·46 (0·29–0·74) | 0·40 (0·26–0·62) | 0·38 (0·27–0·53) |
| 27–29 | 18 | 0·17 (0·09–0·32) | 0·16 (0·06–0·38) | 0·09 (0·03–0·27) | 0·39 (0·26–0·59) | 0·77 (0·40–1·49) | 0·49 (0·31–0·77) | 0·27 (0·18–0·40) |
| 30–32 | 20 | 0·23 (0·17–0·32) | 0·28 (0·11–0·70) | 0·21 (0·14–0·31) | 0·48 (0·28–0·81) | 0·83 (0·50–1·37) | 0·64 (0·41–1·02) | 0·33 (0·25–0·43) |
| 33–35 | 21 | 0·20 (0·16–0·26) | 0·14 (0·06–0·35) | 0·22 (0·16–0·30) | 0·56 (0·43–0·73) | 0·86 (0·58–1·27) | 0·61 (0·42–0·90) | 0·30 (0·23–0·39) |
| 36–38 | 20 | 0·21 (0·16–0·28) | 0·16 (0·09–0·30) | 0·24 (0·13–0·42) | 0·57 (0·39–0·85) | 1·16 (0·66–2·02) | 0·86 (0·54–1·36) | 0·29 (0·24–0·35) |
| 39–41 | 20 | 0·16 (0·10–0·25) | 0·16 (0·07–0·35) | 0·20 (0·12–0·35) | 0·61 (0·37–1·00) | 1·10 (0·62–1·95) | 0·54 (0·34–0·86) | 0·30 (0·19–0·48) |
| 42–44 | 18 | 0·24 (0·17–0·33) | 0·22 (0·12–0·42) | 0·29 (0·16–0·52) | 0·70 (0·45–1·08) | 0·77 (0·43–1·37) | 0·52 (0·31–0·88) | 0·39 (0·26–0·57) |
| 45–47 | 21 | 0·32 (0·19–0·53) | 0·33 (0·14–0·76) | 0·20 (0·13–0·30) | 0·68 (0·47–0·98) | 0·97 (0·68–1·37) | 0·70 (0·41–1·20) | 0·30 (0·23–0·39) |
| 48–50 | 31 | 0·32 (0·23–0·44) | 0·26 (0·15–0·47) | 0·41 (0·24–0·71) | 0·86 (0·61–1·20) | 1·54 (1·12–2·14) | 0·97 (0·68–1·37) | 0·31 (0·22–0·43) |
| 51–53 | 32 | 0·37 (0·28–0·49) | 0·15 (0·07–0·28) | 0·35 (0·22–0·53) | 0·76 (0·53–1·10) | 1·02 (0·73–1·43) | 1·04 (0·75–1·43) | 0·30 (0·22–0·41) |
| 54–56 | 18 | 0·44 (0·29–0·67) | 0·35 (0·14–0·87) | 0·88 (0·49–1·58) | 1·33 (0·90–1·98) | 2·10 (1·51–2·92) | 1·19 (0·87–1·61) | 0·39 (0·26–0·58) |
| 57–59 | 9 | 0·24 (0·10–0·58) | 0·31 (0·14–0·70) | 0·22 (0·10–0·46) | 0·70 (0·24–2·04) | 2·09 (0·44–9·86) | 0·50 (0·27–0·93) | 0·35 (0·20–0·61) |

**Table S4B -** Serotype-specific GMCs calculated using empirical data as shown in manuscript Figure 1, stratified by 3-month age group

| Age band (months) | Serotype 9V | Serotype 14 | Serotype 18C | Serotype 19A | Serotype 19F | Serotype 23F | Serotype 12F | Serotype 33F |
|---|---|---|---|---|---|---|---|---|
| 0–02 | 0·41 (0·18–0·95) | 1·07 (0·42–2·74) | 0·25 (0·11–0·55) | 0·63 (0·32–1·23) | 0·81 (0·29–2·25) | 0·28 (0·10–0·80) | 0·05 (0·01–0·37) | 0·16 (0·06–0·43) |
| 03–05 | 0·96 (0·64–1·45) | 1·50 (0·78–2·86) | 0·78 (0·36–1·70) | 1·71 (1·05–2·79) | 3·50 (2·26–5·43) | 1·09 (0·70–1·70) | 0·15 (0·15–0·15) | 0·14 (0·09–0·21) |
| 06–08 | 0·48 (0·36–0·62) | 1·78 (1·17–2·71) | 0·42 (0·31–0·57) | 1·23 (0·93–1·63) | 1·37 (1·08–1·73) | 0·31 (0·20–0·48) | 0·03 (0·00–0·16) | 0·07 (0·03–0·15) |
| 09–11 | 0·34 (0·26–0·45) | 1·81 (1·36–2·41) | 0·23 (0·17–0·33) | 0·71 (0·53–0·96) | 0·92 (0·66–1·29) | 0·25 (0·19–0·33) | 0·15 (0·15–0·15) | 0·08 (0·04–0·14) |
| 12–14 | 0·35 (0·27–0·45) | 0·90 (0·64–1·25) | 0·18 (0·15–0·23) | 0·88 (0·66–1·17) | 0·95 (0·69–1·31) | 0·21 (0·13–0·34) | 0·02 (0·00–0·99) | 0·09 (0·05–0·18) |
| 15–17 | 0·25 (0·18–0·33) | 1·08 (0·75–1·55) | 0·15 (0·11–0·21) | 0·80 (0·54–1·20) | 0·81 (0·60–1·09) | 0·22 (0·15–0·32) | 0·10 (0·06–0·17) | 0·07 (0·04–0·13) |
| 18–20 | 0·28 (0·19–0·40) | 0·87 (0·59–1·28) | 0·17 (0·11–0·26) | 0·94 (0·61–1·44) | 0·87 (0·60–1·26) | 0·21 (0·12–0·36) | 0·03 (0·01–0·13) | 0·11 (0·06–0·18) |
| 21–23 | 0·34 (0·24–0·48) | 0·85 (0·60–1·19) | 0·14 (0·08–0·23) | 0·89 (0·58–1·38) | 1·20 (0·82–1·77) | 0·20 (0·12–0·35) | 0·06 (0·02–0·16) | 0·13 (0·08–0·21) |
| 24–26 | 0·26 (0·16–0·42) | 0·64 (0·39–1·03) | 0·21 (0·11–0·40) | 0·81 (0·48–1·38) | 1·03 (0·61–1·74) | 0·33 (0·17–0·62) | 0·04 (0·00–0·59) | 0·13 (0·08–0·21) |
| 27–29 | 0·18 (0·09–0·37) | 0·68 (0·35–1·34) | 0·21 (0·10–0·44) | 1·15 (0·65–2·03) | 1·08 (0·59–1·96) | 0·11 (0·03–0·38) | 0·15 (0·15–0·15) | 0·13 (0·05–0·31) |
| 30–32 | 0·32 (0·21–0·49) | 1·88 (1·06–3·31) | 0·19 (0·12–0·29) | 1·25 (0·66–2·39) | 1·52 (0·77–2·99) | 0·23 (0·16–0·33) | 0·03 (0·00–0·37) | 0·13 (0·06–0·30) |
| 33–35 | 0·44 (0·32–0·60) | 1·21 (0·64–2·31) | 0·21 (0·13–0·33) | 2·83 (1·55–5·16) | 1·76 (1·06–2·93) | 0·25 (0·12–0·52) | 0·06 (0·01–0·23) | 0·13 (0·07–0·24) |
| 36–38 | 0·53 (0·27–1·04) | 0·81 (0·39–1·71) | 0·19 (0·09–0·43) | 2·11 (1·37–3·25) | 2·12 (1·18–3·81) | 0·23 (0·12–0·42) | 0·14 (0·10–0·18) | 0·11 (0·04–0·35) |
| 39–41 | 0·32 (0·20–0·52) | 1·25 (0·69–2·28) | 0·25 (0·12–0·50) | 3·04 (1·36–6·77) | 2·18 (1·21–3·92) | 0·21 (0·10–0·47) | 0·05 (0·01–0·30) | 0·18 (0·10–0·30) |
| 42–44 | 0·44 (0·28–0·69) | 1·08 (0·64–1·83) | 0·26 (0·17–0·40) | 1·49 (0·89–2·47) | 1·65 (0·97–2·79) | 0·41 (0·25–0·65) | 0·09 (0·04–0·20) | 0·16 (0·09–0·30) |
| 45–47 | 0·48 (0·35–0·65) | 1·30 (0·77–2·18) | 0·18 (0·12–0·27) | 2·19 (1·36–3·51) | 1·41 (0·90–2·21) | 0·28 (0·13–0·59) | 0·01 (0·00–0·97) | 0·14 (0·08–0·25) |
| 48–50 | 0·81 (0·59–1·12) | 1·12 (0·72–1·75) | 0·41 (0·24–0·68) | 2·44 (1·61–3·71) | 2·45 (1·72–3·49) | 0·49 (0·31–0·78) | 0·04 (0·00–0·26) | 0·19 (0·13–0·27) |
| 51–53 | 0·58 (0·41–0·81) | 1·36 (0·89–2·07) | 0·48 (0·30–0·76) | 2·47 (1·72–3·55) | 2·21 (1·61–3·05) | 0·44 (0·28–0·67) | 0·06 (0·02–0·23) | 0·28 (0·17–0·47) |
| 54–56 | 0·94 (0·65–1·37) | 2·01 (0·98–4·10) | 0·61 (0·36–1·03) | 4·97 (3·08–8·02) | 3·52 (2·24–5·52) | 0·46 (0·31–0·68) | 0·24 (0·11–0·54) | 0·33 (0·21–0·50) |
| 57–59 | 0·66 (0·24–1·79) | 0·94 (0·59–1·51) | 0·28 (0·10–0·74) | 2·84 (1·15–6·99) | 3·57 (0·94,13·65) | 0·28 (0·07–1·10) | 0·32 (0·10–1·02) | 0·58 (0·25–1·37) |

**Table S5 -** Trend among paired (primary and secondary) samples, censored for ages <6 months

| **IgG trend** | **1 % (n)** | **3 % (n)** | **4 % (n)** | **5 % (n)** | **6A % (n)** | **6B % (n)** | **7F % (n)** | **9V % (n)** | **14 % (n)** | **18C % (n)** | **19A % (n)** | **19F % (n)** | **23F % (n)** | **12F % (n)** | **33F % (n)** |
|---|---|---|---|---|---|---|---|---|---|---|---|---|---|---|---|
| Increase ≥4-fold[¥] | 4·1 (3) | 8·5 (6) | 5·2 (4) | 4·0 (3) | 2·6 (2) | 5·2 (2) | 1·3 (1) | 5·5 (4) | 6·6 (5) | 4·2 (3) | 10·5 (8) | 4·0 (3) | 8·2 (6) | 0·0 (0) | 2·6 (2) |
| Increase ≥2-fold[¶] | 12·5 (9) | 17·1 (12) | 15·7 (12) | 10·8 (8) | 15·7 (12) | 10·5 (8) | 4· 0(3) | 15·2 (11) | 10·6 (8) | 11·2 (8) | 18·4 (14) | 13·5 (10) | 16·4 (12) | 3·1 (2) | 9·3 (7) |
| Increase <2-fold | 6·9 (5) | 18·5 (13) | 11·8 (9) | 37·8 (28) | 22·3 (17) | 21·0(16) | 16·0 (12) | 15·2 (11) | 9·3 (7) | 9·8 (7) | 27·6 (21) | 25·6 (19) | 10·9 (8) | 1·5 (1) | 9·3 (7) |
| Any increase, % | 19·4 | 35·6 | 27·5 | 48·6 | 38·0 | 31·5 | 20·0 | 30·4 | 19·9 | 21·0 | 46·0 | 39·1 | 27·3 | 4·6 | 18·6 |
| No change | 0·0 (0) | 0·0 (0) | 0·0 (0) | 0·0 (0) | 0·0 (0) | 0·0 (0) | 0·0 (0) | 0·0 (0) | 1·3 (1) | 1·4 (1) | 0·0 (0) | 0·0 (0) | 0·0 (0) | 0·0 (0) | 0·0 (0) |
| LLD[§] at V1 & V2 | 12·5 (9) | 17·1 (12) | 25·0 (19) | 5·4 (4) | 5·2 (4) | 5·2 (4) | 4·0 (3) | 16·6 (12) | 1·3 (1) | 23·9 (17) | 1·3 (1) | 0·0 (0) | 31·5 (23) | 84·3 (54) | 48·0 (36) |
| Decrease | 68·0 (49) | 47·1 (33) | 47·3 (36) | 45·9 (34) | 56·5 (43) | 63·1 (48) | 76·0 (57) | 52·7 (38) | 77·3 (58) | 53·5 (38) | 52·6 (40) | 60·8 (45) | 41·1 (30) | 10·9 (7) | 33·3 (25) |
| Total pairs in analysis, N[¥] | 72 | 70 | 76 | 74 | 76 | 76 | 75 | 72 | 75 | 71 | 76 | 74 | 73 | 64 | 75 |

[§]LLD = lower limit of detection of assay (0·15µg/mL). VT=vaccine serotype, NVT=non-vaccine serotype

[¶]Increase ≥2-fold includes all samples with increase ≥4-fold; [¥] Increase ≥4-fold may include all or a subset of those with increase ≥2-fold.

[¥]Though 82 pairs were collected and sent for laboratory analysis, some samples had results that excluded them from this analysis (incl., inadequate volume, no IgG detected).

Table S5 reports the trend in paired samples, showing either a decrease, an increase or no change in IgG titre when comparing primary and secondary samples. An increase in IgG titre is evidence of a novel pneumococcal acquisition event, we hypothesise after the primary sample and before the secondary sample, resulting in an increase in serotype-specific IgG. An alternative explanation for an increase in IgG is a novel acquisition event occurring before the primary sample but with an increased IgG (relative to the primary sample) still detected at secondary sample. A decrease in IgG titre is evidence of no novel pneumococcal acquisition event after the primary sample. Samples with LLD at both visits had detectable IgG too low to quantify, reported as 0.075µg/mL (LLD*0.5) for both primary and secondary samples.

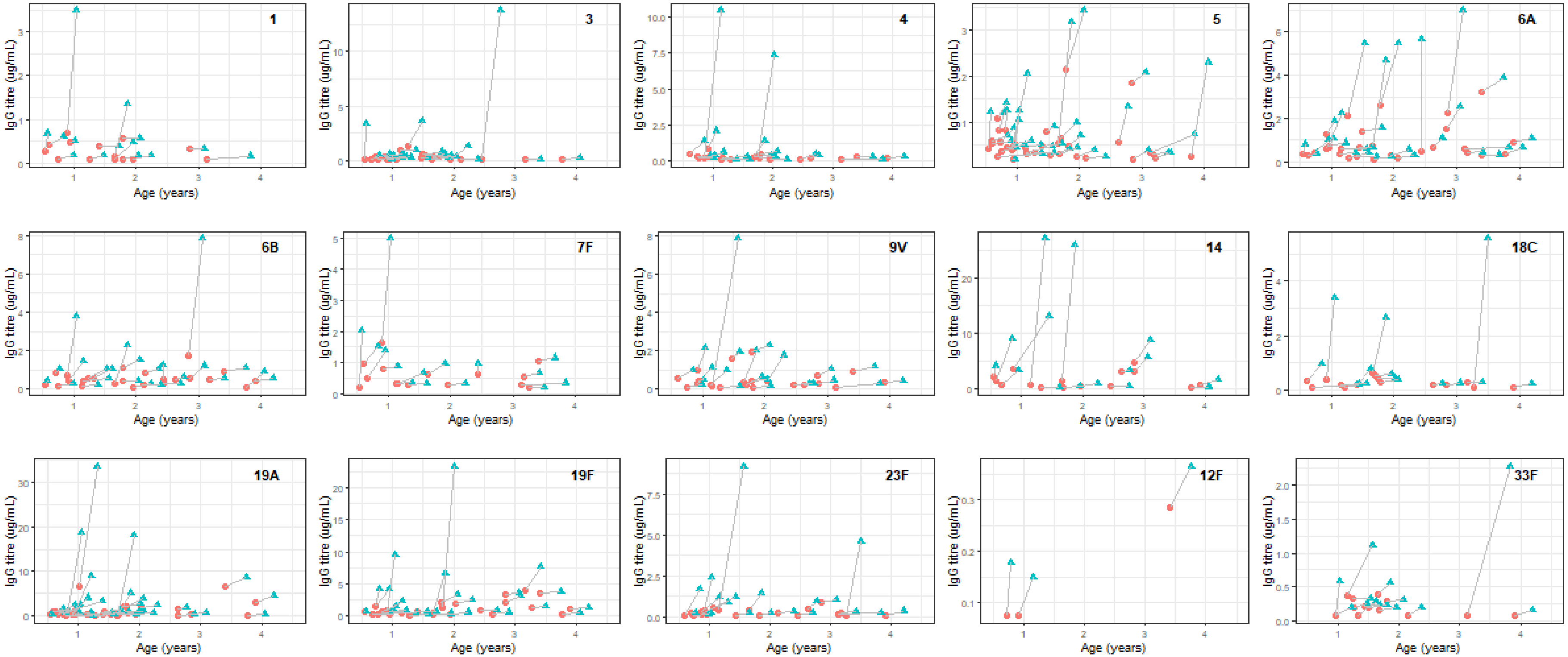


**Figure S1 -** Seroincident events, based on 82 linked pairs

Seroincident events based on 82 linked pairs with the respective secondary serum samples collected approximately 3 months after the primary samples. Data reported are limited to pairs with any increase in IgG titres (positive slope). Red dot=primary sample. Blue triangle=secondary sample.

**2A. PsaA**

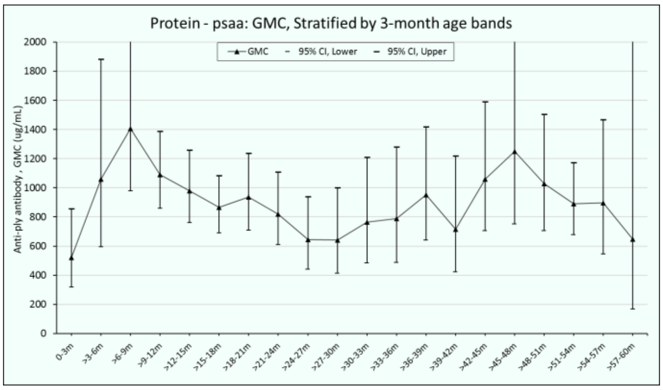


**2B. NanA**

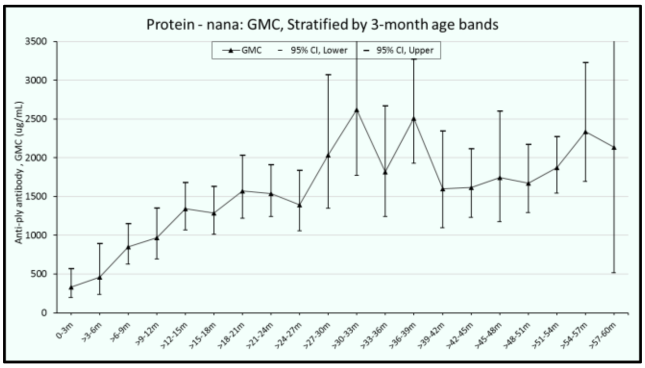


**2C. Ply**

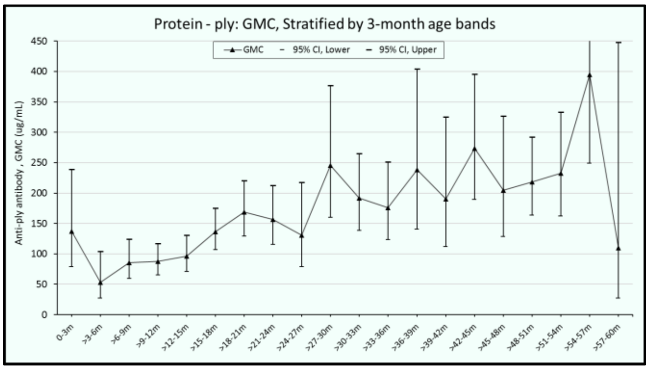


**Figure S2 -** Protein-specific antibody profiles, stratified by 3-month age groups

**Table S6 -** Median IgG titre and carriage prevalence (rounds 4–6 in PCVPA Study) in the 3 to 6 months and 6 to 12 months age groups

| | 3-6 months | | 6-12 months | |
|---|---|---|---|---|
| | **IgG titre, median n=19** | **Carriage prevalence (%) n=37** | **IgG titre, median n=113** | **Carriage prevalence (%) n=117** |
| Lower transmission | | | | |
| 4 | 1·27 | 0 (0%) | 0·35 | 0 (0%) |
| 6B | 0·46 | 0 (0%) | 0·58 | 0 (0%) |
| 7F | 1·20 | 0 (0%) | 0·79 | 0 (0%) |
| 9V | 0·78 | 0 (0%) | 0·38 | 1 (0·85%) |
| 18C | 1·02 | 0 (0%) | 0·30 | 0 (0%) |
| Higher transmission | | | | |
| 6A | 0·83 | 1 (2·7%) | 0·63 | 5 (4·2%) |
| 14 | 1·42 | 1 (2·7%) | 2·19 | 6 (5·1%) |
| 19A | 1·90 | 1 (2·7%) | 0·90 | 2 (1·7%) |
| 19F | 3·20 | 0 (0%) | 0·92 | 4 (3·4%) |
| 23F | 1·07 | 1 (2·7%) | 0·23 | 2 (1·7%) |

**Table S7.** Percent of children attaining CoP thresholds (μg/mL) post vaccination, stratified by age group

| Age | Serotype 1 (n= 534) | | | Serotype 3 (n=519) | | | Serotype 4 (n=554) | | | Serotype 5 (n=553) | | |
|---|---|---|---|---|---|---|---|---|---|---|---|---|
| | ≥ 0·35[¥] | ≥ CoPd[¶] | ≥ CoPc[§] | ≥ 0.35[¥] | ≥ CoPd[¶] | ≥ CoPc[§] | ≥ 0·35[¥] | ≥ CoPd[¶] | ≥ CoPc[§] | ≥ 0·35[¥] | ≥ CoPd[¶] | ≥ CoPc[§] |
| **<3m** | 58·3 | 50·0 | 50·0 | 41·6 | 16·6 | - | 58·3 | - | 25·0 | 58·3 | - | 41·6 |
| **3-5m** | 87·5 | 81·3 | 81·3 | 73·3 | 6·6 | - | 75·0 | - | 56·2 | 87·5 | - | 56·2 |
| **6-11 m** | 67·4 | 28·1 | 25·8 | 40·0 | 1·1 | - | 51·6 | - | 12·0 | 61·1 | - | 31·1 |
| **1y** | 20·3 | 3·4 | 2·8 | 36·9 | 3·4 | - | 23·6 | - | 2·1 | 49·7 | - | 23·2 |
| **2y** | 26·5 | 4·8 | 4·8 | 36·2 | 7·5 | - | 25·0 | - | 2·3 | 58·3 | - | 30·9 |
| **3y** | 22·7 | 6·7 | 6·7 | 33·8 | 2·8 | - | 36·0 | - | 10·6 | 69·7 | - | 40·7 |
| **4y** | 48·8 | 13·4 | 13·4 | 35·0 | 3·7 | - | 56·6 | - | 25·5 | 81·1 | - | 55·5 |
| **Total** | 36·7 | 13·1 | 12·6 | 37·7 | 4·1 | - | 37·7 | - | 10·8 | 62·0 | - | 34·7 |

| Age | Serotype 6A (n=555) | | | Serotype 6B (n=552) | | | Serotype 7F (n=553) | | | Serotype 9V (n=546) | | |
|---|---|---|---|---|---|---|---|---|---|---|---|---|
| | ≥ 0·35[¥] | ≥ CoPd[¶] | ≥ CoPc[§] | ≥ 0·35[¥] | ≥ CoPd[¶] | ≥ CoPc[§] | ≥ 0·35[¥] | ≥ CoPd[¶] | ≥ CoPc[§] | ≥ 0·35[¥] | ≥ CoPd[¶] | ≥ CoPc[§] |
| **<3m** | 41·6 | 66·6 | - | 25·0 | 66·6 | 25·0 | 41·6 | 33·3 | 16·6 | 45·4 | 36.3 | 18.1 |
| **3-5m** | 87·5 | 93·7 | - | 68·7 | 87·5 | 43·7 | 81·2 | 68·7 | 56·2 | 93·7 | 75.0 | 43.7 |
| **6-11 m** | 73·6 | 92·3 | - | 73·6 | 92·3 | 60·4 | 85·7 | 50·5 | 25·2 | 56·6 | 25.5 | 10.0 |
| **1y** | 59·6 | 90·8 | - | 54·6 | 86·8 | 35·5 | 62·1 | 21·0 | 10·8 | 43·6 | 22.6 | 8.2 |
| **2y** | 73·8 | 95·2 | - | 69·0 | 96·4 | 44·0 | 46·4 | 8·3 | 3·5 | 46·3 | 19.5 | 6.1 |
| **3y** | 80·2 | 90·7 | - | 72·3 | 86·8 | 56·5 | 36·8 | 10·5 | 2·6 | 55·2 | 32.8 | 15.7 |
| **4y** | 92·2 | 100 | - | 85·6 | 97·7 | 75·5 | 46·0 | 13·4 | 4·4 | 78·8 | 60.0 | 24.4 |
| **Total** | 72·6 | 92·7 | - | 67·2 | 90·5 | 50·3 | 57·6 | 22·9 | 11·3 | 55·1 | 32.0 | 13.1 |

| Age | Serotype 14 (n= 554) | | | Serotype 18C (n= 546) | | | Serotype 19A (n=555) | | | Serotype 19F (n=552) | | |
|---|---|---|---|---|---|---|---|---|---|---|---|---|
| **<3m** | 75·0 | 66·6 | 33·3 | 25·0 | 75·0 | 16·6 | 66·6 | 66·6 | - | 83·3 | 25.0 | 25.0 |
| **3-5m** | 87·5 | 81·2 | 43·7 | 68·7 | 81·2 | 50·0 | 100·0 | 100 | - | 100 | 93. | 68.7 |
| **6-11 m** | 86·8 | 82·4 | 43·9 | 49·4 | 71·9 | 8·9 | 80·2 | 8·9 | - | 92·1 | 39.3 | 13.4 |
| **1y** | 79·0 | 73·1 | 17·2 | 20·2 | 52·4 | 3·2 | 73·6 | 8·4 | - | 77·9 | 36.0 | 18.8 |
| **2y** | 77·3 | 66·6 | 25·0 | 26·5 | 65·0 | 8·4 | 82·1 | 82·1 | - | 86·7 | 55.4 | 26.5 |
| **3y** | 82·6 | 73·3 | 26·6 | 32·8 | 63·0 | 5·4 | 89·4 | 89·4 | - | 92·1 | 71.0 | 36.8 |
| **4y** | 87·7 | 81·1 | 25·5 | 57·7 | 78·8 | 16·6 | 96·6 | 96·6 | - | 98·8 | 78.8 | 45.5 |
| **Total** | 82·1 | 75·0 | 26·5 | 35·3 | 64·6 | 9·1 | 82·5 | 82·5 | - | 87·6 | 52.7 | 27.5 |

| Age | Serotype 23F (n=554) | | | Serotype 12F (n=455) | | | Serotype 33F (n=455) | | |
|---|---|---|---|---|---|---|---|---|---|
| **<3m** | 50.0 | 66.6 | 41.6 | 8·3 | - | - | 8·3 | - | - |
| **3-5m** | 87.5 | 100.0 | 68.7 | 0·0 | - | - | 0·0 | - | - |
| **6-11 m** | 35.9 | 60.6 | 21.3 | 1·1 | - | - | 1·1 | - | - |
| **1y** | 36.2 | 48.6 | 21.6 | 1·1 | - | - | 1·1 | - | - |
| **2y** | 37.3 | 55.4 | 22.8 | 5·9 | - | - | 5·9 | - | - |
| **3y** | 40.5 | 58.1 | 27.0 | 1·8 | - | - | 1·8 | - | - |
| **4y** | 60.2 | 73.8 | 36.3 | 19·2 | - | - | 19·2 | - | - |
| **Total** | 42.6 | 58.8 | 26.6 | 4·1 | - | - | 4·1 | - | - |

[¥]Aggregate CoP for IPD (Siber and colleagues)[7], [¶]Putative CoP for disease (Andrews and colleagues)[8]: serotype 1=0·78, 3=2·83, 4=0·35, 5=undefined, 6A=0·16, 6B=0·16, 7F=0·87, 9V=0·62, 14=0·46, 18C=0·14, 19A=1·00, 19F=1·17, 23F=0·20; [§]Putative correlate of protection for carriage (Voysey and colleagues)[11]: serotype 1=0·81, 3=undefined, 4=1·16, 5=0·73; 6A=undefined, 6B=0·50, 7F=1·60, 9V=1·31, 14=2·48, 18C=1·32, 19A=undefined, 19F=2·54, 23F=0·63. Dash (-) in cell indicates that the CoP is undefined. m=month, y=year

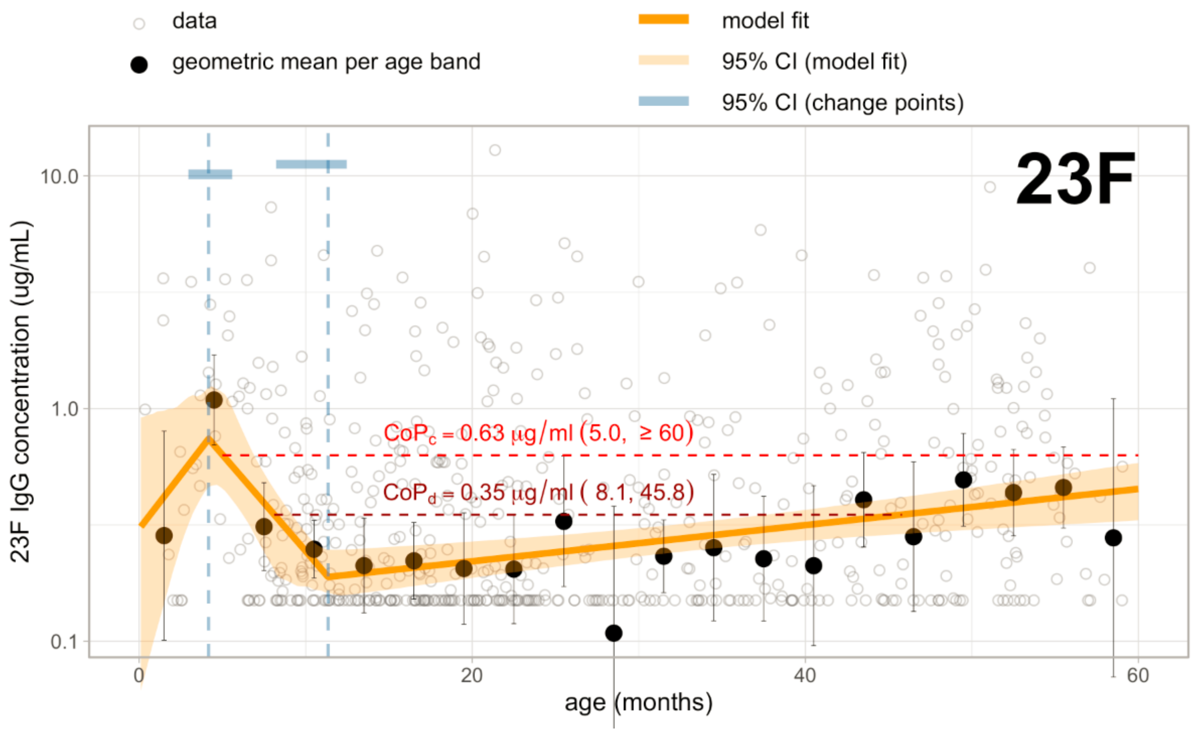


**Figure S3 -** Population anti-23F titre trends, age periods when above or below CoP

Figure S3 is the spline regression model output for VT 23F (as shown in figure 1 of the main manuscript) with age periods indicated when population level IgG falls below CoP. Small hollow dots indicate empirical IgG titre datapoints for each sample. Black dots are the geometric mean concentrations (GMCs) for all datapoints within 3-month age bands. The orange line is the spline model fit and the orange shaded area is the 95% CI for the model fit. Vertical dashed lines indicate changepoints. Blue bars at the top of each image are the 95% CI for the age at each change point.

In this figure, anti-23F antibody titres are estimated to be below CoP for carriage from ages 5·0 to ≥60 months (ie, beyond the age included in this analysis) and below the CoP for IPD from the age 8·1 to 45.78 months. Refer to Table 4 in the manuscript for estimated age bands with CoPs below the putative thresholds for disease and carriage for other key pneumococcal serotypes.

$CoP_c$=CoP threshold for protection against pneumococcal carriage (serotype-specific as reported by Voysey at al.[3]). $CoP_d$=correlate of protection threshold for protection against invasive pneumococcal disease (aggregate threshold as reported by Siber at al.[1]); Horizontal red dashed line indicates when population anti-23F antibody titres are below the CoP. Horizontal blue dashed lines show when population anti-23F antibody titres are above the CoP.
